# Soluble immune checkpoints are dysregulated in COVID-19 and heavy alcohol users with HIV infection

**DOI:** 10.1101/2021.12.22.21268218

**Authors:** Wei Li, Fahim Syed, Richard Yu, Jing Yang, Ying Xia, Ryan F. Relich, Shanxiang Zhang, Mandana Khalili, Laurence Huang, Melissa A. Kacena, Xiaoqun Zheng, Qigui Yu

## Abstract

Immune checkpoints (ICPs) consist of paired receptor-ligand molecules that exert inhibitory or stimulatory effects on immune defense, surveillance, regulation, and self-tolerance. ICPs exist in both membrane and soluble forms *in vivo* and *in vitro*. Imbalances between inhibitory and stimulatory membrane-bound ICPs (mICPs) in malignant cells and immune cells in the tumor immune microenvironment (TIME) have been well documented. Blockades of inhibitory mICPs have emerged as an immense breakthrough in cancer therapeutics. However, the origin, structure, production regulation, and biological significance of soluble ICPs (sICPs) in health and disease largely remains elusive. Soluble ICPs can be generated through either alternative mRNA splicing and secretion or protease-mediated shedding from mICPs. Since sICPs are found in the bloodstream, they likely form a circulating immune regulatory system. In fact, there is increasing evidence that sICPs exhibit biological functions including (1) regulation of antibacterial immunity, (2) interaction with their mICP compartments to positively or negatively regulate immune responses, and (3) competition with their mICP compartments for binding to the ICP blocking antibodies, thereby reducing the efficacy of ICP blockade therapies. Here, we summarize current data of sICPs in cancer and infectious diseases. We particularly focus on sICPs in COVID-19 and HIV infection as they are the two ongoing global pandemics and have created the world’s most serious public health challenges. A “storm” of sICPs occurs in the peripheral circulation of COVID-19 patients and is associated with the severity of COVID-19. Similarly, sICPs are highly dysregulated in people living with HIV (PLHIV) and some sICPs remain dysregulated in PLHIV on antiretroviral therapy (ART), indicating these sICPs may serve as biomarkers of incomplete immune reconstitution in PLHIV on ART. We reveal that HIV infection in the setting of alcohol abuse exacerbates sICP dysregulation as PLHIV with heavy alcohol consumption have significantly elevated plasma levels of many sICPs. Thus, both stimulatory and inhibitory sICPs are present in the bloodstream of healthy people and their balance can be disrupted under pathophysiological conditions such as cancer, COVID-19, HIV infection, and alcohol abuse. There is an urgent need to study the role of sICPs in immune regulation in health and disease.

## Introduction

Immune checkpoints (ICPs) consist of paired receptor-ligand molecules that exert inhibitory or stimulatory effects on immune defense, surveillance, regulation, and self-tolerance^1-5^. Under normal circumstances, ICPs regulate the breadth, magnitude, and duration of the immune responses against malignancy and infection while protecting tissues from excessive insult. However, in certain pathological situations such as cancer or persistent infection, the balance between ICP stimulatory and inhibitory signals becomes dysregulated^1-7^. Malignant cells can dysregulate the expression of ICPs on the surface of immune cells to evade or subvert the immune response, leading to insufficiency or failure of anti-tumor immune attacks. The up-regulated expression of inhibitory ICPs, including CTLA-4, PD-1, TIM-3, and LAG-3 has been found on the surface of both CD4 and CD8 T cells in cancer patients^6,8-10^. These important findings have laid the foundation for the clinical development of ICP blockade therapies, which abrogate ICP inhibitory signals, restoring and enhancing the anti-tumor activity of cytotoxic T lymphocytes (CTLs)^3,5,11,12^. Since 2011, ICP blockers targeting CTLA-4, PD-1, and PD-L1 have yielded unprecedented responses in a portion of cancer patients, leading to seven FDA-approved ICP blocking antibodies against these ICPs, including one anti-CTLA-4 antibody (Ipilimumab), three anti-PD-1 antibodies (Nivolumab, Pembrolizumab, and Cemiplimab), and three anti-PD-L1 antibodies (Avelumab, Durvalumab, and Atezolizumab), for treating several types of cancer such as melanoma and lung cancer^5,13-15^.

Similar to malignant cells, several pathogens, including HIV (the human immunodeficiency virus), HBV (hepatitis B virus), TB (tuberculosis), and malaria have been demonstrated to dysregulate ICPs to limit host-protective CTLs^6,7,16,17^. For example, in antiretroviral therapy (ART)-naïve people living with HIV (PLHIV), there is upregulated expression of multiple inhibitory ICPs including CTLA-4, PD-1, TIM-3, and LAG-3 on total and HIV-specific CD4 and CD8 T cells^6-8,17-21^, which is associated with an accelerated decline in the number of CD4 T cells in PLHIV^8,22^. Following ART, expression of these ICPs on the surface of T cells declines, but remains elevated when compared with healthy controls^8-10^. Importantly, expression of inhibitory ICPs can be used as surrogate immunological biomarkers of ART effectiveness. In ART-treated PLHIV, PD-1 expression on CD8 T cells has been associated with impaired reconstitution of CD4 T cells and a shorter time to viral rebound after stopping ART^23,24^. In addition, CD4 T cells expressing high levels of PD-1, LAG-3, and TIGIT alone or in combination are enriched for integrated HIV DNA during ART^25,26^. Furthermore, most CD4 T cells expressing at least one of these ICPs, carry inducible and replication-competent HIV genomes^25^. Thus, CD4 T cells expressing inhibitory ICPs such as PD-1 and LAG-3 contribute to HIV persistence during ART. *In vitro* and *ex vivo* studies have shown that blocking antibodies against either PD-1, CTLA-4, LAG-3, or TIM-3 can significantly restore the proliferative capacities and functions of T cells and B cells from PLHIV on ART^21^. Currently, there are several clinical trials in PLHIV with and without cancer using the FDA-approved ICP blockers^6,21,27-31^. These clinical trials have yielded mixed results ranging from little therapeutic benefit to significant expansions of HIV-specific CD4 and CD8 T cells in a subset of participants^27-32^, indicating ICP blockade for the treatment of HIV infection needs further study.

ICP molecules exist in both membrane and soluble forms *in vivo* and *in vitro*^33-44^. Similar to membrane-bound ICPs (mICPs), soluble ICPs (sICPs) are also present in normal physiological conditions and highly dysregulated in patients with cancer, viral infections, or ALD (alcohol-associated liver disease)^33-45^. Soluble ICPs can be generated through either alternative mRNA splicing and secretion or protease-mediated shedding from mICPs by actions of matrix metalloproteinases (MMPs)^44,46^. Although the detailed structure, production regulation, function, and clinical relevance of sICPs largely remain unknown, many sICPs exist in native forms that exhibit biological functions such as regulation of antibacterial immunity^42,46^. Since sICPs are paired receptor-ligand molecules and circulate in the bloodstream, they likely form a circulating immune regulatory system. In addition, increasing evidence has shown that sICPs interact with their mICP compartments to positively or negatively regulate immune responses^43^. Furthermore, sICPs can compete with their mICP compartments for binding to the ICP blocking antibodies, thereby interrupting the efficacy of ICP blockade therapies. Thus, there is an urgent need to study the role of sICPs in immune regulation in health and disease. Here, we summarize current data of sICPs in cancer and infectious diseases. We particularly focus on sICPs in COVID-19 and HIV infection as they are the two ongoing global pandemics and have created the world’s most serious public health and development challenges.

### Soluble immune checkpoints in the peripheral circulation of healthy people

As described in our recent report^45^, we used a multiplex immunoassay (the Human Immuno-Oncology Checkpoint Protein Panel, MilliporeSigma, Burlington, MA) to simultaneously quantify plasma concentrations of 16 sICPs (sBTLA, sCD27, sCD28, sCD40, sCD80/B7-1, sCD86/B7-2, sCTLA-4, sGITR, sGITRL, sHVEM, sICOS, sLAG-3, sPD-1, sPD-L1, sTIM-3, and sTLR-2) in healthy blood donors. Plasma levels of two additional ICPs (sCD160 and sLIGHT) were quantified using the Human CD160 Matched ELISA Antibody Pair Set (Sino Biological, Beijing, China) and the Human LIGHT Duoset ELISA Kit (R&D Systems, Minneapolis, MN), respectively. We found that, except for sGITR and sLIGHT, which were only detected in 30-50% of plasma samples, all other 16 sICPs were steadily detected in plasma samples from all healthy blood donors (Table 1). These sICPs could be generated through either alternative mRNA splicing and secretion or protease-mediated shedding from mICPs by actions of MMPs^44,46^. Several studies have demonstrated that sICPs are present in the peripheral blood as the native polypeptide products of their genes and have biological functions. For example, CTLA-4, also known as CD152, is a member of the immunoglobulin (Ig) gene superfamily^47^. CTLA4 is constitutively expressed in regulatory T cells but can be upregulated in conventional T cells after activation^47,48^. CTLA-4 and CD28 are homologous receptors that share a pair of ligands (B7-1 and B7-2) expressed on the surface of antigen-presenting cells (APCs), but mediate opposing functions in T-cell activation^48^. CTLA-4 interacts with its ligands to inhibit T-cell responses^48^, while CD28 acts as a major co-stimulatory receptor in promoting full activation of T cells in response to T cell receptor (TCR) engagement^49^. As shown in Table 1, plasma levels of sCTLA-4 in healthy individuals were detected at a median concentration of 31 pg/mL with the interquartile ranges of 12 – 81 pg/mL (n=39), which are higher than the detection limit (9.3 pg/mL) of the multiplex immunoassay. These results argue against a previous report showing that circulating CTLA-4 was undetectable in healthy volunteers using an enzyme immunoassay (EIA)^44^. This is because the EIA has limited detection sensitivity (≥ 4 ng/mL)^44^, which is insufficiently sensitive for the detection of sCTLA-4 concentrations in healthy people. Immunoprecipitation and Western blotting analyses of serum sCTLA-4 revealed a polypeptide consistent with the predicted size (23 kDa) from an alternative transcript of the *CTLA-4* gene, suggesting sCTLA-4 is present as a native molecule rather than a product of proteolytic digestion or shedding of mCTLA-4^44^. Functional studies have shown that sCTLA-4 immunoreactivity can be blocked by B7.1 (also known as CD80), one of its known ligands. This supports the notion that sCTLA-4 is present as a soluble functional molecule. Thus, sCTLA-4 likely has important immunoregulatory functions, which is similar to soluble cytokine receptors such as soluble forms of TNF receptor, IL-2α receptor, IL-4 receptor, and IL-7 receptor that exist in the biological fluids and regulate cytokine activity *in vitro* and *in vivo*^50-55^.

**Table 1.** Plasma levels of sICPs in healthy adults.

| **Stimulatory ICPs** | **Healthy adults (n=34-39)** | **Sensitivity (pg/mL)** | **Inhibitory ICPs** | **Healthy adults (n=34-39)** | **Sensitivity (pg/mL)** |
| --- | --- | --- | --- | --- | --- |
| **CD27** | 881<br>(603-1,180) | 24.1 | BTLA | 370<br>(139-668) | 43.8 |
| **CD28** | 108<br>(496-2,106) | 84.5 | CD160 | 5,590<br>(3,801-10,605) | 93.8 |
| **CD40** | 285<br>(148-385) | 4.3 | CTLA-4 | 31<br>(12-81) | 9.3 |
| **CD80** | 21<br>(13-31) | 11.2 | HVEM | 1,197<br>(885-1,651) | 0.8 |
| **CD86** | 718<br>(457-1,228) | 86.1 | LAG-3 | 3,459<br>(2,120-4,852) | 66.0 |
| **GITR** | 16<br>(6-48) | 18.8 | PD-1 | 361<br>(219-744) | 13.7 |
| **GITRL** | 169<br>(81-427) | 20.5 | PD-L1 | 19<br>(10-41) | 1.3 |
| **ICOS** | 370<br>(139-668) | 55.6 | TIM-3 | 1,228<br>(1,021-1,875) | 1.5 |
| **LIGHT** | 240<br>(39-1,274) | 62.5 |  |  |  |
| **TLR-2** | 466<br>(286-935) | 24.1 |  |  |  |

**Note:** Data are represented as median and (interquartile ranges) in pg/mL. Characteristics of study subjects were described in our previous report<sup>45</sup>.

In contrast to sCTLA-4, several sICPs including sHVEM (also known as TNFRSF14 or CD270), sCD160, sLAG-3, and sTIM-3 were present at high concentrations (Table 1) in healthy people. These are potent inhibitory ICPs. HVEM was initially identified as the receptor of herpes simplex virus 1 (HSV-1) through binding to the HSV-1 glycoprotein D (gD)^56^. Since then, HVEM has been identified as a co-signaling molecular switch through interacting with BTLA (also known as CD272), CD160, and LIGHT^57^. In addition, HVEM can bind to SALM5 (synaptic adhesion-like molecule 5) to regulate neuroinflammation^58^. We have recently reported that recombinant sHVEM affects TNF-α and IFN-γ production by anti-CD3/anti-CD28-stimulated T cells from healthy volunteers^45^, indicating sHVEM may act as a circulating immune regulator.

Like sCTLA-4 and sHVEM, other sICPs such as sLAG-3, sTIM-3, sPD-1, and sPD-L1 are biologically active and participate in immune regulation^39,59-61^. Thus, the majority (if not all) of ICPs have soluble forms that are detectable in the peripheral blood of healthy individuals. Different sICPs are likely produced at different levels and at distinct checkpoints to fine-tune immune homeostasis in health, although their origin, production regulation, and biological function are yet to be discovered. Of note, these data related to plasma sICPs were obtained from adults with a median age of 42 years (26-52) and 64% were from males^45^. Future study is needed to investigate whether sICP levels and functions in healthy people are affected by age, gender, and race.

### Soluble immune checkpoints in cancer

ICPs act as gatekeepers for immune responses and play a central role in immune homeostasis that is maintained by a precise balance between stimulatory and inhibitory ICPs on the surface of effector and regulatory immune cells. The immune homeostasis is a tightly regulated network which fails during tumor development due to an imbalance between inhibitory and stimulatory ICPs. Indeed, high levels of inhibitory ICPs on the surface of tumor cells is a hallmark of the tumor immune microenvironment (TIME) that is infiltrated with many types of innate and adaptive immune cells^62^. Increased inhibitory ICPs are responsible for tumor immune escape and thereby have become major targets for cancer immunotherapy^5,13-15^.

While mICPs have been extensively studied in cancer immunity and cancer immunotherapy, the origin, production regulation, and biological significance of sICPs largely remains elusive. Due to their function in both positive and negative immune regulation, sICPs and their levels change in the peripheral blood, which may affect the development, prognosis, and treatment of cancer. Studies have shown that plasma or serum levels of sICPs can serve as biomarkers and/or predictors of cancer patient outcomes or therapeutic responses^43,63^. Plasma or serum levels of numerous sICPs including sPD-L1, sPD-1, sLAG-3, sTIM-3, sCTLA-4, sHVEM, sCD80, sCD86, sCD27, sCD40, and sBTLA are highly elevated in patients with various types of tumors and serve as prognostic markers for solid tumors such as non-small cell lung cancer, gastric cancer, colon cancer, and cervical cancer^42,60,64-67^. Soluble ICPs are also biologically active in cancer patients. Studies have revealed that plasma/serum levels of sCD40L are highly elevated in patients with lung cancer and undifferentiated nasopharyngeal carcinoma^68,69^. The elevated sCD40L in cancer patients is likely derived from activated platelets rather than T cells, because cancer patients have significant platelet activation, but inadequate T-cell activation^70-72^. A functional study has shown that the upregulated sCD40L seen in cancer patients exerts an immunosuppressive effect through enhancement of MDSC (myeloid-derived suppressor cell)-mediated suppression of T cell proliferation and IFN-γ production, expansion of regulatory T cells (Treg), and enrichment of PD-1^+^ T cells^73^. On the other hand, sPD-1 is likely generated through mRNA splicing and secretion, as four PD-1 splice variants have been identified^74^. *In vitro* and *in vivo* studies have shown sPD-1 is able to bind its membrane-bound ligands (mPD-L1 and mPD-L2) to block mPD-1/mPD-L1/mPD-L2 interaction, thereby restoring T cell immunity^60^. Indeed, local delivery of sPD-1 in the tumor microenvironment through adeno-associated virus-mediated delivery vector induces antitumor immunity through improving T cell function^75,76^. The origin, production regulation, function, and biological significance of sICPs in tumors have been systematically reviewed^43,60,77,78^. Collectively, the levels of sICPs in the peripheral circulation in cancer patient are frequently altered, which likely has clinical significances. That said, a better understanding of the underlying mechanisms of the sICP network could lay the foundation for the development of new strategies for treating cancers with immunotherapies.

### Soluble immune checkpoints in patients with COVID-19

Two recent studies, reported by Kong Y *et al*. (2020) and Avendano-Ortiz J *et al*. (2021), have demonstrated that a “storm” of sICPs occurs in COVID-19 patients and is associated with the severity of COVID-19^79,80^. The Kong study quantified 14 sICPs including sBTLA, sCTLA-4, sGITR, sHVEM, sIDO, sLAG-3, sPD-1, sPD-L1, sPD-L2, sTIM-3, sCD27, sCD28, sCD80, and s4-1BB in the serum samples from patients with asymptomatic, mild/moderate, and severe/critical COVID-19 using the ProcartaPlex Human ImmunoOncology Checkpoint Panel (Invitrogen, Carlsbad, CA)^79^, while the Avendano-Ortiz study quantified 9 sICPs including sCD25, sCD86, sCTLA-4, Galectin-9, sLAG-3, sPD-1, sPD-L1, sTim-3, and s4-1BB using the LEGENDplex HU Immune Checkpoint Panel 1 (BioLegend, San Diego, CA)^80^. After merging the overlapping 6 sICPs that were detected in both studies, a total of 17 sICPs including sBTLA, sCTLA-4, sGalectin-9, sGITR, sHVEM, sIDO, sLAG-3, sPD-1, sPD-L1, sPD-L2, sTIM-3, sCD25, sCD27, sCD28, sCD80, sCD86, and s4-1BB were studied in the serum or plasma samples from COVID-19 patients^79,80^. The Kong study showed that, except for sPD-L2, each of the other 13 sICPs was significantly higher in the severe/critical group than in the mild/moderate and asymptomatic groups^79^. On the other hand, the Avendano-Ortiz study showed that plasma levels of sCD25, sCD86, Galectin-9, sPD-1, sPD-L1, and sTim-3, but not sLAG-3, sCTLA-4, and s4-1BB, were significantly higher in the severe/critical group than in the mild/moderate groups^80^. Therefore, both studies demonstrated that the serum or plasma levels of sPD-1, sPD-L1, and sTIM-3 were significantly higher in the severe/critical group than in the mild/moderate and asymptomatic groups, but their data conflicted regarding the serum or plasma levels of sLAG-3, sCTLA-4, and s4-1BB between healthy controls and COVID-19 patients^80^. The Kong study also showed that serum levels of 11 sICPs (sGITR, s4-1BB, sTIM-3, sCD27, sLAG-3, sPD-1, sCD28, sCTLA-4, sBTLA, sHVEM, and sCD80) were persistently higher in severe/critical patients than in mild/moderate cases during hospitalization. In addition, the levels of 8 sICPs (sIDO, sGITR, s4-1BB, sTIM-3, sCD27, sLAG-3, sPD-1, and sCD28) were negatively correlated with absolute counts of CD4 and CD8 T cells. The Avendano-Ortiz study also demonstrated that plasma levels of sCD25, sTIM-3, Galectin-9, and sPD-L1, but not sCD86, showed a negative correlation with the absolute lymphocyte count (ALC). These results suggest that sICPs are dysregulated in COVID-19 and sICP dysregulation may be linked to COVID-19 lymphopenia, an abnormal reduction in lymphocyte numbers. Lymphopenia is a prominent clinical feature of COVID-19 patients and has been associated with the severity of COVID-19^81-86^. Indeed, non-survivors of COVID-19 have a significantly lower lymphocyte count than survivors^82,87^. The absolute cell counts of lymphoid lineage cells, including T cells, B cells, and NK cells, are abnormally reduced with a more pronounced decrease in CD8 T cells^88-90^. In contrast, myeloid lineage cells such as neutrophils are highly increased in the blood of patients with severe COVID-19^83^, which is noted as a major clinical feature of severe COVID-19^91^. The mechanisms of COVID-19 lymphopenia remain unclear, although several hypotheses are proposed including a cytokine storm impact^92-94^, direct infection of immune cells^95^, overaggressive T cell responses^96^, and lymphocyte infiltration and sequestration in the lungs^87^. However, these hypotheses have been challenged, because ***(1)*** most COVID-19 patients do not have remarkably high levels of inflammatory cytokines, as only 4% of critically ill COVID-19 patients develop cytokine storm symptom (CSS) and anti-CSS medications have no benefit for most COVID-19 patients^83,97-101^, ***(2)*** direct viral infection is an unlikely cause of immune cell loss^102^, as infectious SARS-CoV-2 has not been successfully isolated from peripheral blood cells in COVID-19 patients^94^, ***(3)*** the overall magnitude of the T cell response in COVID-19 patients is either insufficient or excessive remains debated^96^, as T cell responses are insufficient in some COVID-19 patients, but excessive in others^96^, and ***(4)*** post-mortem biopsies from COVID-19 patients with marked lymphopenia reveal prominent infiltration of neutrophils, but neither T cells nor B cells, in the lungs^103-106^. Thus, studies are urgently needed to determine the cause and impact of the commonly observed lymphopenia in patients with severe COVID-19, and whether dysregulated sICPs are associated with the pathogenesis of COVID-19 lymphopenia.

We also used a multiplex immunoassay (the Human Immuno-Oncology Checkpoint Protein Panel, MilliporeSigma, Burlington, MA) to simultaneously quantify the concentrations of 16 sICPs in plasma samples from healthy controls (n=23) and patients with asymptomatic (n=15) or hospitalized (severe/critical) COVID-19 (n=24). Among these 16 sICPs, 4 (sCD40, sGITRL, sICOS, and sTLR-2) were not previously studied in COVID-19 patients, while 12 were already tested in the two studies by Kong and Avendano-Ortiz *et al*. In our studies, the healthy control subjects were matched with the COVID-19 patients in terms of age, sex, and race. We found that plasma levels of the majority of 16 sICPs were significantly higher in COVID-19 patients when compared with healthy controls (data not shown). The 4 sICPs (sCD40, sGITRL, sICOS, and sTLR-2) that had not previously been studied in COVID-19 were significantly higher in the plasma from patients with asymptomatic or severe/critical COVID-19 when compared with healthy controls (Fig. 1A). Plasma levels of sGITRL, sICOS, and sTLR-2, but not sCD40 were further elevated in severe/critical COVID-19 patients than in asymptomatic cases (Fig. 1A). We also found that sCTLA-4 and sLAG-3, 2 sICPs that were studied by Kong and Avendano-Ortiz *et al*. with conflicting results, were elevated in plasma from patients with severe/critical COVID-19 when compared with healthy controls (Fig. 1B), which is in agreement with Kong’s results.

**Figure 1.**
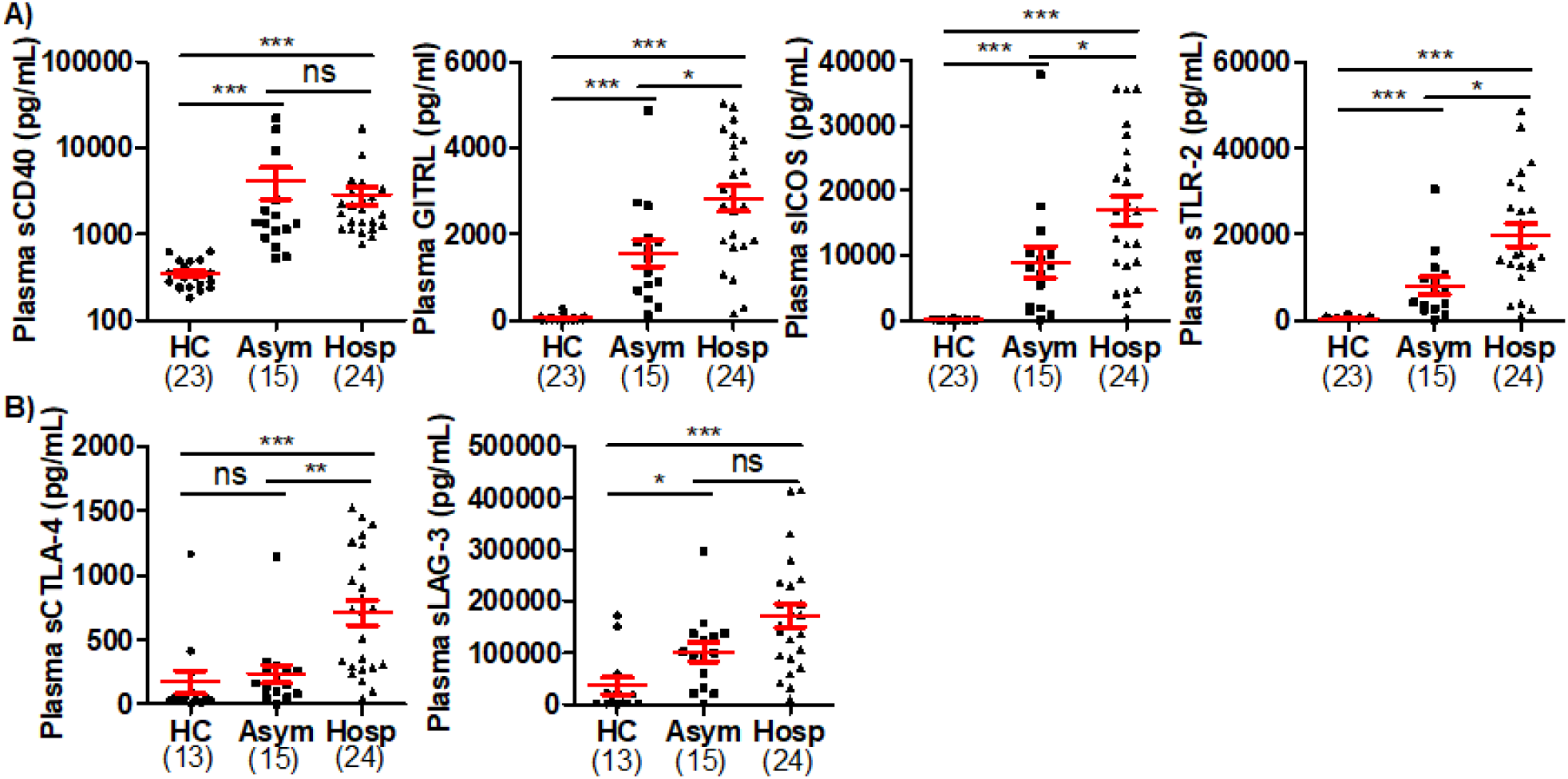
Plasma levels of sICPs were highly elevated in COVID-19. **A)** Scatter plots demonstrating the plasma levels of 4 sICPs that were not previously studied in COVID-19. **B)** Scatter plots demonstrating the plasma levels of 2 sICPs in COVID-19 that were previously reported with different results. Kruskal-Wallis test with Dunn’s corrections for pairwise comparisons among hospitalized (severe/critical) COVID-19 patients (Hosp), SARS-CoV-2-infected individuals without symptoms (Asym), and healthy controls (HCs). Red lines represent the mean and the standard error of the mean. ns, no significant; \**p*<0.05; \*\**p*<0.01; \*\*\**p*<0.001.

Taken together, a storm of sICPs occurs in the peripheral circulation of COVID-19 patients and is associated with the disease severity. The circulating sICP levels on admission appear to be better mortality predictors than inflammatory cytokines and chemokines^80^, and thereby can potentially serve as biomarkers of COVID-19 progress and outcome. Given that some, if not all, sICPs are biological active, they may also serve as circulating immune regulators or pharmaceutical targets for COVID-19 therapy. To this end, mechanistic studies and large-scale, cross-sectional and longitudinal studies are needed to investigate the origin, production regulation, and clinical significance of sICPs in patients with COVID-19.

### Soluble immune checkpoints in people living with HIV (PLHIV)

Chronic immune activation and exhaustion are important features of persistent viral infections such as infection with HIV. Indeed, immune exhaustion represents a barrier to effective and specific immunity against HIV infection. Chronic immune activation and exhaustion are at least in part attributed to the dysregulation of ICPs. In addition to mICPs that have been demonstrated to play a critical role in immune homeostasis in PLHIV, sICPs may also be dysregulated in PLHIV and thereby contribute to immune exhaustion in PLHIV. A recent study used ELISA to quantify plasma levels of sPD-L1 in PLHIV and healthy controls and found that plasma levels of sPD-L1 were significantly elevated in PLHIV and remained high despite control of HIV infection by ART^107^. In addition, PLHIV on ART with virological failure had the highest plasma levels of sPD-L1^107^. Thus, sPD-L1 in the peripheral blood represents a potential biomarker of immune exhaustion and virological failure in PLHIV.

Here, we simultaneously quantified plasma levels of 16 sICPs from 23 healthy controls, 46 PLHIV who were ART-naïve, and 65 PLHIV who were on ART using a multiplex immunoassay as detailed above. These three groups of study subjects were matched in terms of demographic parameters including age, gender, and race. As shown in Fig. 2, except GITR which was only detected in 34.8% (8/23) of healthy controls, each of the other 15 sICPs was detectable in the plasma samples from healthy controls, indicating that they could play biological roles in immune homeostasis under physiologic conditions. Compared to healthy controls, ART-naïve PLHIV had significantly higher plasma levels of the 15 sICPs tested. Specifically, only sTIM-3 was not elevated in ART-naïve PLHIV. In comparison to healthy controls, PLHIV on ART only had higher plasma levels of 3 sICPs (sCD40, sCTLA-4, and sHVEM) (Fig. 2). These findings appear to indicate that ART effectively, but not completely, restores ICP homeostasis. Among these 3 sICPs that remained at higher levels in the peripheral blood of PLHIV on ART, sHVEM and sCD40 were not affected by ART, while sCTLA-4 was dramatically reduced, but did not return to a normal level (Fig. 2). Both CD40 (also known as TNFRSF5) and HVEM (also known as TNFRSF14) are members of the tumor necrosis factor receptor superfamily. CD40 is a co-stimulatory molecule that is mainly expressed on the surface of APCs such as dendritic cells, monocytes/macrophages, and B cells. CD40 is required for APC activation via binding to CD154 (also known as CD40 ligand or CD40L) on T cells. CD40-CD40L interaction leads to the initiation of bidirectional intracellular signaling in both CD40^+^ APCs and CD40L^+^ T cells, resulting in APC activation and T cell responses^108-110^. Dysregulation of CD40/CD40L expression and interactions contributes to the severity in numerous diseases such as HIV infection^111-113^, cancer^108,114^, and autoimmune disorders^115-117^. However, the role of sCD40 in the peripheral circulation of PLHIV largely remains elusive. We found that ART-naïve PLHIV had excessive production of sCD40, which was minimally affected by ART, suggesting that circulating sCD40 may represent an indicator of dysregulation of APC and T cell function that is a hallmark of HIV-associated deficiency in cell-mediated immunity.

**Figure 2.**
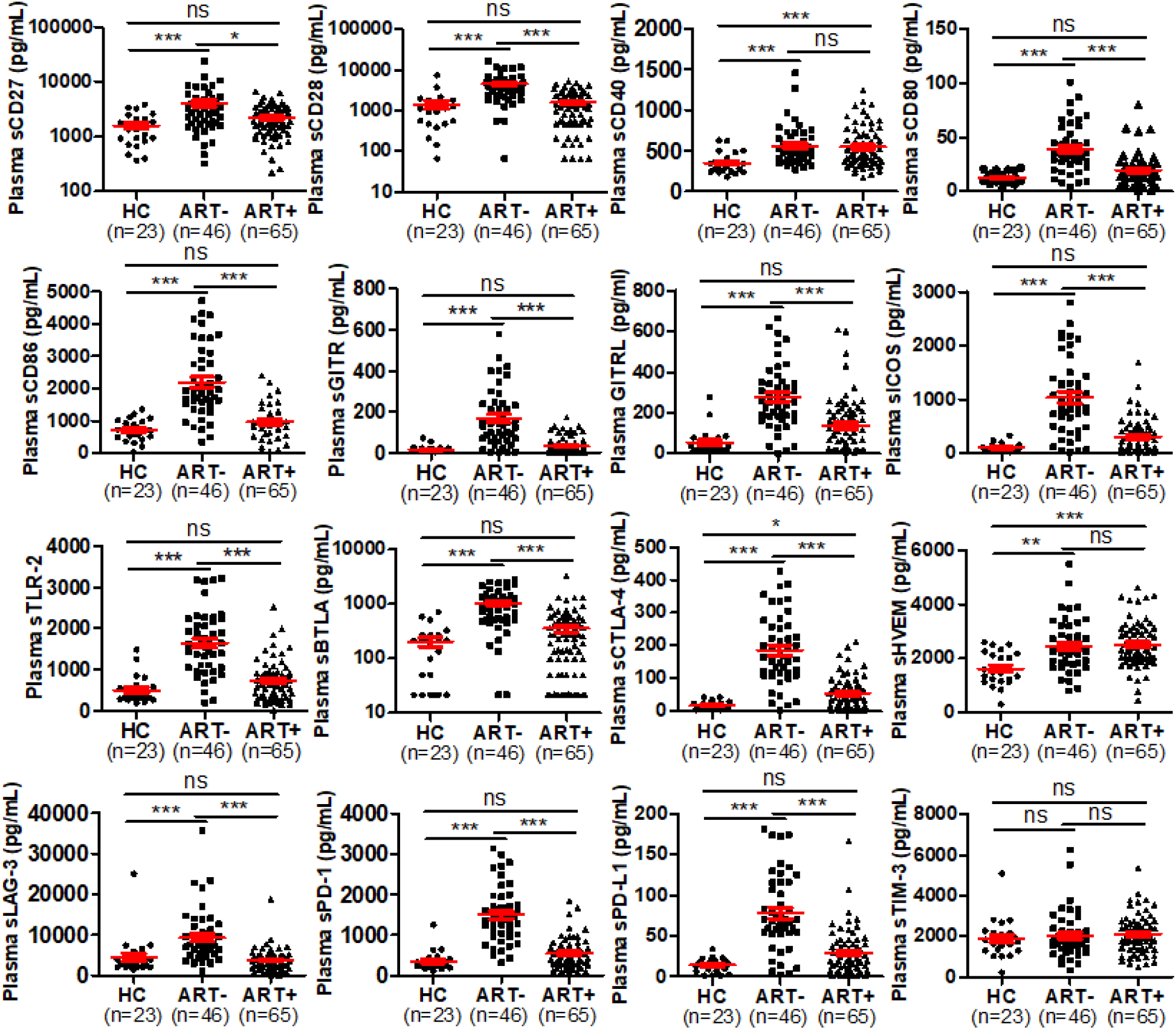
Plasma levels of sICPs were highly dysregulated in PLHIV. Scatter plots demonstrating the plasma levels of sICPs in HCs (healthy controls), ART-naïve PLHIV, and PLHIV on ART. Kruskal-Wallis test with Dunn’s corrections for pairwise comparisons among PLHIV on ART, ART-naïve PLHIV, and HCs. Red lines represent the mean and the standard error of the mean. ns, no significant; \**p*<0.05; \*\**p*<0.01; \*\*\**p*<0.001. Abbreviations: HC, healthy control; ART-, people living with HIV (PLHIV) who were not treated with antiretroviral therapy (ART); and ART+, people living with HIV (PLHIV) who were treated with antiretroviral therapy (ART).

Similar to sCD40, sHVEM was also excessively produced in PLHIV and was barely affected by ART (Fig. 2). HVEM serves as a shared receptor/ligand for stimulatory and inhibitory ligands/receptors, including LIGHT, BTLA, and CD160 that are expressed on both hematopoietic and non-hematopoietic cells. HVEM acts as a bifunctional ligand/receptor that exhibits costimulatory signals upon binding to LIGHT and co-inhibitory signals upon binding to BTLA or CD160^118,119^. Due to its role of bifunctional ligand/receptor, HVEM serves as a molecular switch between stimulatory and inhibitory signaling^120,121^, thereby playing a unique role in immune homeostasis. We have recently reported that expressions of both sHVEM and mHVEM were highly dysregulated in heavy alcohol users with ALD, specifically alcoholic hepatitis (AH), when compared with heavy alcohol users without AH^45^. Plasma levels of upregulated sHVEM in AH patients remained high for 6 months of complete alcohol abstinence^45^, indicating sHVEM might serve as a prognostic marker for AH. We also found that sHVEM-his, consisting of the soluble extracellular domain of human mHVEM linked to a polyhistidine tag at the C-terminus, significantly inhibited TCR-induced TNF-α production by both CD4 T cells and CD8 T cells from AH patients and healthy controls^45^, indicating sHVEM plays an inhibitory role in HVEM axis-mediated TNF-α production. Currently, the regulation and function of sHVEM in HIV immunopathogenesis is not known. Thus, studies are needed to elucidate the mechanisms of action for the sHVEM axis and the interplay between sHVEM and mHVEM in HIV infection.

### Soluble immune checkpoints in heavy alcohol users with HIV infection

Alcohol abuse and HIV infection are both major health issues worldwide. Globally, more than 2 billion people consume alcohol on a regular basis, and approximately 76 million suffer from alcohol-related disorders^122,123^. Long-term heavy alcohol users develop a spectrum of ALD, ranging from AH, fibrosis/cirrhosis, to hepatocellular carcinoma (HCC)^124^. Alcohol overconsumption contributes to 5.1% of the global burden of diseases and causes approximately 3.3 million deaths every year^125,126^. HIV is the causative agent of AIDS and has claimed over 36 million lives with an estimated 38 million PLHIV worldwide at the end of 2020^127^. Alcohol overconsumption is common among PLHIV and adversely influences the health outcomes by increasing HIV-associated comorbidities such as liver disease, cardiovascular disease, pulmonary disease, bone disease, and cancer^128-130^. It is well known that alcohol overconsumption and HIV infection independently damage the gastrointestinal (GI) tract mucosal barrier, leading to a leaky gut that allows microbial translocation and accumulation of microbial components such as lipopolysaccharide (LPS) in the blood. Specifically, alcohol disrupts gap junction integrity of gut mucosal epithelial cells, leading to increased GI permeability and translocation of microbial components such as LPS from the GI tract into the blood and liver^131-133^. Alcohol-induced microbial translocation has been considered a major driver of chronic immune activation and inflammation in AH patients^134-137^. In PLHIV, microbial translocation is a cause of chronic immune activation and inflammation, which is a hallmark of progressive HIV infection and a stronger predictor of disease outcome compared to plasma viral load^138,139^. We therefore hypothesize that alcohol overconsumption and HIV infection exacerbate microbial translocation, immune dysregulation, and inflammation, thereby accelerating disease progression of HIV infection and ALD. To test this hypothesis, we established a cohort of heavy alcohol users with and without HIV infection. We analyzed and compared the profiles of sICPs in the peripheral blood in heavy drinkers without overt ALD (HDC) versus PLHIV on ART (HIV) versus PLHIV on ART who were heavy drinkers, but did not have ALD (HDC+HIV). We found that plasma levels of all 16 sICPs were similar between HDC and HIV groups (Table 2). Fourteen out of sixteen sICP examined were dramatically elevated in the peripheral blood in HDC+HIV when compared with either HDC or HIV (Table 2). sCD27 and sTIM-3 were not elevated in HDC+HIV compared with either HDC or HIV (Table 2). These results indicate that sICPs were highly dysregulated in HDC+HIV even though these individuals had no clinical evidence of overt ALD and their liver enzyme parameters including the circulating levels of AST and ALT and AST:ALT ratio were similar to healthy individuals (data not shown). Previous studies from our group and others have demonstrated that chronic excessive drinking leads to immune abnormalities in heavy drinkers even when there are no obvious signs of clinical liver disease^137,140-142^. HDCs have increased bacterial translocation as they have higher serum levels of LPS and markers of monocyte/macrophage activation (sCD14 and sCD163) than non-excessive drinkers^141^. In addition, mucosal-associated invariant T (MAIT) cells in the peripheral blood are significantly decreased in HDCs compared to healthy controls^142^. Moreover, HDCs have increased levels of MAIT activation-associated cytokines such as IL-18 and IL-12^142^. MAIT cells are innate-like lymphocytes that are highly enriched in liver, mucosa, and peripheral blood, and play a protective role in antimicrobial immunity^143,144^. These results highlight the presence of immune dysregulation in HDCs. HIV infection and alcohol abuse dramatically exacerbate immune abnormalities such as sICP dysregulation. Future study is needed to investigate the mechanisms underlying exacerbated sICP dysregulation in heavy drinkers with HIV infection.

**Table 2.** Plasma levels of sICPs in heavy alcohol users with HIV infection.

| sICP | HDC (n=26) | HIV (n=28) | HDC+HIV (n=21) | P value |
| --- | --- | --- | --- | --- |
| sBTLA | 2,89 (126-533) | 336 (203-453) | 650 (407-948) | <sup>ns</sup> *P* <sub>1</sub>, *** *P* <sub>2</sub>, ** *P* <sub>3</sub> |
| sCD27 | 1,375 (746-1,855) | 1,701 (697-3,244) | 1,727 (805-4,214) | <sup>ns</sup>*P*<sub>1</sub>, <sup>ns</sup>*P*<sub>2</sub>, <sup>ns</sup>*P*<sub>3</sub> |
| sCD28 | 1,201 (341-2,481) | 1,254 (665-1,979) | 3,043 (1,796-5,934) | <sup>ns</sup> *P* <sub>1</sub>, ** *P* <sub>2</sub>, ** *P* <sub>3</sub> |
| sTIM-3 | 1,789 (1,240-2,526) | 2,023 (1,084-2,348) | 2,715 (1,529-3,339) | <sup>ns</sup>*P*<sub>1</sub>, <sup>ns</sup>*P*<sub>2</sub>, <sup>ns</sup>*P*<sub>3</sub> |
| sHVEM | 1,809 (762-2,385) | 2,208 (1,980-2,808) | 3,723 (2,459-5,148) | <sup>ns</sup> *P* <sub>1</sub>, *** *P* <sub>2</sub>, * *P* <sub>3</sub> |
| sCD40 | 589 (158 – 706) | 643 (439-815) | 1,207 (909-1,519) | <sup>ns</sup> *P* <sub>1</sub>, *** *P* <sub>2</sub>, *** *P* <sub>3</sub> |
| sLAG-3 | 3,138 (1,136-5,776) | 3,932 (1,688-5,632) | 7,282 (4,232-8,709) | <sup>ns</sup> *P* <sub>1</sub>, ** *P* <sub>2</sub>, ** *P* <sub>3</sub> |
| sTLR-2 | 579 (44-165) | 788 (554-1,027) | 1,437 (1,067-1,739) | <sup>ns</sup> *P* <sub>1</sub>, *** *P* <sub>2</sub>, *** *P* <sub>3</sub> |
| sGITRL | 123 (140-905) | 120 (85-178) | 245 (163-345) | <sup>ns</sup> *P* <sub>1</sub>, *** *P* <sub>2</sub>, *** *P* <sub>3</sub> |
| sPD-1 | 647 (201-980) | 675 (467-1,148) | 1,962 (1,206-2,290) | <sup>ns</sup> *P* <sub>1</sub>, *** *P* <sub>2</sub>, *** *P* <sub>3</sub> |
| sCTLA-4 | 78 (16-124) | 56 (19-99) | 250 (142-386) | <sup>ns</sup> *P* <sub>1</sub>, *** *P* <sub>2</sub>, *** *P* <sub>3</sub> |
| sCD80 | 30 (17-45) | 34 (18-45) | 51 (28-77) | <sup>ns</sup> *P* <sub>1</sub>, ** *P* <sub>2</sub>, * *P* <sub>3</sub> |
| sCD86 | 870 (222-1,619) | 912 (673-1,585) | 2,521 (1,355-3,747) | <sup>ns</sup> *P* <sub>1</sub>, *** *P* <sub>2</sub>, ** *P* <sub>3</sub> |
| sPD-L1 | 30 (10-51) | 29 (20-48) | 65 (46-90) | <sup>ns</sup> *P* <sub>1</sub>, *** *P* <sub>2</sub>, *** *P* <sub>3</sub> |
| sGITR | 86 (0-291) | 21 (0-137) | 696 (332-935) | <sup>ns</sup> *P* <sub>1</sub>, *** *P* <sub>2</sub>, *** *P* <sub>3</sub> |
| sICOS | 166 (52-296) | 205 (134-269) | 340 (242-450) | <sup>ns</sup> *P* <sub>1</sub>, *** *P* <sub>2</sub>, *** *P* <sub>3</sub> |

**Note:** Data are represented as median (interquartile range). Kruskal-Wallis test with Dunn's
corrections was used to calculate differences among 3 groups of HDCs, HIV, and HDC+HIV.
HDCs, heavy alcohol drinkers without overt liver disease; HIV, people living with HIV (PLHIV) on antiviral therapy (ART); HDC+HIV, HDCs with HIV infection on ART, but without overt liver
disease. *P*<sub>1</sub>, statistical analysis between HDC and HIV; *P*<sub>2</sub>, statistical analysis between HDC and HDV+HIV; *P*<sub>3</sub>, statistical analysis between HIV and HDC+HIV. <sup>ns</sup>*P*, no significant; \**P*<0.05;
\*\**P*<0.01; \*\*\**P*<0.001. Detailed definitions of HDC and the inclusion and exclusion criteria were previously described<sup>137,145</sup>.

## Conclusion

ICP molecules exist in both membrane and soluble forms *in vivo* and *in vitro*. Imbalance between inhibitory and stimulatory mICPs in malignant cells and immune cells in tumor microenvironment has been well documented and blockade of inhibitory mICPs has emerged as an immense breakthrough in cancer therapeutics. While mICPs have been extensively studied, their soluble compartments have not been adequately studied. sICPs can be generated through either alternative mRNA splicing and secretion or protease-mediated shedding from mICPs. However, the cellular resource, structure, production regulation, and biological significance of sICPs in health and disease largely remain elusive. Here, we summarize current data of sICPs in cancer and infectious diseases. A storm of sICPs occurs in the peripheral circulation of COVID-19 patients and is associated with the severity of COVID-19. Similarly, sICPs are highly dysregulated in PLHIV and some sICPs remain dysregulated in PLHIV on ART, indicating these sICPs may serve as biomarkers of incomplete immune reconstitution for PLHIV on ART. Strikingly, HIV infection and alcohol abuse dramatically exacerbate sICP dysregulation. Thus, both stimulatory and inhibitory sICPs are present in the bloodstream of healthy people and their balance can be disrupted under pathophysiological conditions such as cancer, COVID-19, HIV infection, and alcohol abuse. Further studies are needed to investigate whether sICPs act as critical circulating immune regulators in health and disease.

## Data Availability

All data produced in the present study are available upon reasonable request to the authors

## Abbreviations

ALD: alcohol-associated liver disease
APC: antigen-presenting cell
ART: antiretroviral therapy
BTLA: B- and T-lymphocyte attenuator
CTLA-4: cytotoxic T-lymphocyte-associated protein 4
GITR: glucocorticoid-induced tumor necrosis factor receptor-related protein
GITRL: glucocorticoid-induced tumor necrosis factor receptor-ligand
HBV: hepatitis B virus
HC: healthy control
HCC: hepatocellular carcinoma
HIV: human immunodeficiency virus
HVEM: herpesvirus entry mediator
ICP: immune checkpoint
ICOS: inducible T-cell costimulator
LAG-3: lymphocyte-activation gene 3
LIGHT: homologous to lymphotoxin, exhibits inducible expression and competes with HSV glycoprotein D for binding to herpesvirus entry mediator, a receptor expressed on T lymphocytes
mICP: membrane-bound immune checkpoint
PD-1: programmed death 1
PD-L1: programmed death-ligand 1
PLHIV: people living with HIV
sICP: soluble immune checkpoint
TB: tuberculosis
TCR: T cell receptor
TIM-3: T-cell immunoglobulin and mucin domain 3
TIME: tumor immune microenvironment
TNFRSF: tumor necrosis factor receptor superfamily
TLR-2: Toll-like receptor-2

## Notes

### Study Subjects and Ethical Considerations

This study was performed with the approval of the Institutional Review Board at Indiana University School of Medicine and University of California San Francisco. Blood samples were drawn after each participant provided a written informed consent form.

#### Disclaimer

The content of this article is solely the responsibility of the authors and does not necessarily represent the official views of the National Institutes of Health (NIH) or the Bill and Melinda Gates Foundation.

#### Financial support

This work was supported by the National Institute on Alcohol Abuse and Alcoholism (grant number UH2AA026218 to Q. Y.); the Bill and Melinda Gates Foundation (grant number OPP1035237 to Q. Y.); the Indiana Biobank and the Indiana Clinical and Translational Sciences Institute funded by the NIH (grant number UL1TR002529); and the National Center for Advancing Translational Sciences, Clinical and Translational Sciences Award. Dr. Khalili was also in part supported by the National Institute of Alcohol Abuse and Alcoholism (grant number K24AA022523).

